# Tocilizumab Effect in COVID-19 Hospitalized Patients: A Systematic Review and Meta-Analysis of Randomized Control Trials

**DOI:** 10.1101/2021.03.15.21253581

**Authors:** Basheer Abdulrahman, Waleed Aletreby, Ahmed Mady, Alfateh Mohammed Noor, Mohammed Lhmdi, Fahad Faqihi, Abdulrahman Alharthy, Mohammed Al-Odat, Dimitrios Karakitsos, Ziad Memish

## Abstract

Since the emergence of the first cases of COVID-19 viral pneumonia late 2019 several studies evaluated the benefits of different treatment modalities. Early in the pandemic, the interleukin 6 (IL-6) receptor antibody Tocilizumab was considered in view of the cytokine release syndrome associated with COVID-19 infection. Several early observational studies showed beneficial effect of treatment with Tocilizumab on mortality, however, results from well-designed randomized clinical trials (RCT) were contradicting.

**Objectives:** To perform a systematic literature review and meta-analysis of RCTs utilizing Tocilizumab in the treatment of COVID-19 pneumonia, with in-hospital mortality as a primary objective, while secondary objectives included composite outcome of mortality, intubation, or ICU admission, another secondary outcome was super added infection.

**Method:** This was a random effects model (DerSimonian and Laird) model of relative risk (RR), along with corresponding 95% confidence intervals, p values, and forest plots of both primary and secondary outcomes. A fixed effect sensitivity test was performed for the primary outcome, in addition to subgroup and meta-regression analyses with predefined criteria.

**Results:** The primary outcome of mortality showed statistically insignificant reduction of mortality with Tocilizumab (RR = 0.9, 95% CI: 0.8 – 1.01; p = 0.09) although with an unmistakable apparent clinical benefit. There was a significant reduction in the RR of the secondary composite outcome (RR = 0.83, 95% CI: 0.76 – 0.9; p < 0.001), and no difference between groups in super-added infection (RR = 0.77, 95% CI: 0.51 – 1.19; p = 0.24). Treatment protocol allowing a second dose was the only significant predictor of improved mortality in the meta-regression analysis. Certainty of evidence was reduced to moderate for the primary outcome and the secondary outcome of clinical deterioration, while it was reduced to low for the secondary outcome of super-added infection.

**Conclusion:** Moderate certainty of evidence suggest no statistically significant improvement of 28-30 day all-cause mortality of hospitalized COVID-19 patients treated with TCZ, although there may be clinically important value. Moderate certainty of evidence suggest lowered relative risk of a composite outcome of death or clinical deterioration, while, low grade evidence indicate no increase in the risk of super-added infection associated with TCZ treatment. A protocol allowing two doses of TCZ shows evidence of improved mortality as compared to a strictly single dose protocol.

## Introduction

Since the first case of severe acute respiratory syndrome coronavirus 2 (COVID-19) infection was identified at the end of 2019, COVID-19 has become a huge threat to global health ^[1,2]^. The full spectrum of clinical manifestations of COVID-19 ranges from asymptomatic carriage and mild acute respiratory disease, to severe pneumonia and even acute respiratory distress syndrome (ARDS) ^[3]^. Although, different mortality reports were coming, most of the deaths were attributed to severe COVID-19 cases ^[4]^. COVID-19 is a novel emerging infectious disease associated with a complicated pathogenesis; however, laboratory evidence of severe COVID-19 infections suggests that cytokine release syndrome (CRS) plays a crucial pathogenic role ^[5]^. Although many proinflammatory cytokines are involved in CRS, interleukin-6 (IL-6) is the most important, although it was also found to be a poor prognostic factor ^[6]^.

Tocilizumab (TCZ) is a humanized monoclonal antibody that can target both membrane-bound and soluble forms of the IL-6 receptor, and several studies have evaluated its efficacy in the treatment of severe COVID-19, tocilizumab use showed a rapid and sustained response and was also associated with significant clinical improvement. By neutralizing a key inflammatory factor in the cytokine release syndrome (CRS), this molecule may block the cytokine storm during the systemic hyper-inflammation stage and reduce disease severity ^[7,8]^. In another study by Ramaswamy et al., although tocilizumab-treated patients displayed higher levels of biomarkers [C-reactive protein (CRP) and IL-6] indicative of cytokine storm at initial presentation, tocilizumab still provided a short-term survival benefit ^[9]^, such results of observational studies were also reflected in systematic reviews ^[10]^. However, contradicting results are emerging from well-designed randomized clinical trials (RCT), indicating lack of such benefit ^[11]^, in opposition to other RCTs clearly reflecting clinical and mortality benefits^[12]^.

In view of the conflicting evidence, and the increasing number of published RCTs we aimed to conduct the current systematic literature review to try and fill in the gap of evidence, and consider it as an update of previous reviews that included only observational studies, or those that included both observational and randomized trials.

## Objectives

The primary objective of the review was to compare 28 – 30 day all-cause mortality among hospitalized patients with confirmed COVID-19 infection whom were given TCZ, to the mortality of similar control patients.

Secondary objectives included the comparison of the same groups with regards to: Combined outcome of death, intubation, or intensive care unit (ICU) admission, and incidence of super-infection.

## Method

We utilized the PRISMA checklist of preferred reporting items for systematic reviews and meta-analysis.

### Study selection criteria

We included only RCTs that compared head to head at least two arms, one receiving TCZ (as intervention arm) and another not receiving TCZ (as the control), if TCZ was being compared to another medication, that medication’s group was considered the control, but if the study included more than two arms, the TCZ arm was compared to the control only, without consideration of the third arm, regardless of TCZ dosing regimen.

The included studies must have recruited adult patients (at least 18 years old) with confirmed COVID-19 infection, regardless of other inclusion or exclusion criteria pertaining to the severity of the condition (such as oxygen requirement), or pre-specified laboratory tests’ values. Included studies must have also reported at least one of the objectives of this review.

### Search strategy

We systematically searched for eligible studies in PUBMED, EMBASE, and medical archives (medRxiv) using the keywords: “COVID-19”, “SARS-CoV-2”, and “Tocilizumab” (details of Pubmed search in supplementary file). Furthermore, we reviewed the list of references of each potentially eligible article for additional studies. The final search list was reviewed by three authors (AB, MA, AW) for final inclusion in the review, any disagreements were resolved by discussion.

### Risk of bias (RoB) and quality of evidence assessment

Each included study was evaluated independently by 2 authors for RoB using the modified version of the Cochrane Collaboration Tool ^[13]^, the tool is built in within the Review Manager ® software. The Cochrane Collaboration tool assesses RoB in each included study with regards to 7 domains, namely: random sequence generation, allocation concealment, blinding of participants, blinding of assessors, attrition bias, selective reporting bias, in addition to other sources of bias. Each one of the 7 domains can be evaluated on a 3 level scale as low, unclear, or high risk of bias. The RoB evaluation of each study as well as a summary RoB of included studies were graphically presented.

As for the quality of evidence, we evaluated each outcome according to the GRADE methodology ^[14]^, very briefly the methodology evaluates certainty of evidence for a particular outcome after the consideration of 5 criteria: Individual study risk of bias, directness, consistency, precision, and publication bias. Accordingly, generated evidence of each outcome can be graded as: high, moderate, low, or very low.

### Data abstracting

Each included study was abstracted twice by two independent authors for comparison and validation of consistency. Each author summarized an included study on a pre-prepared excel sheet that included the following information: Last name of first author, year of publication, country, number of patients in intervention and control groups, as well as total number of patients. Data abstracting also included details of inclusion and exclusion criteria, dosing regimen of TCZ, and a list of reported outcome.

### Publication bias assessment

We assessed publication bias of the primary outcome using Egger’s test (considered significant for publication bias if p value < 0.05), according to the test result we presented a trim-and-fill funnel plot.

### Statistical method

Both the primary and secondary objectives of this review were dichotomous outcomes, accordingly, were presented as risk ratio (RR), with corresponding 95% confidence interval (CI). In view of our anticipation of existing between studies differences, at least in terms of studied populations and TCZ dosing, we used DerSimonian and Laird method in a random effects model to pool the effect size of each outcome, and presented corresponding forest plots, along with corresponding 95% CI and p values. For each reported outcome we evaluated heterogeneity using I^2^ test, and considered heterogeneity among included studies to be high if I^2^ was higher than 50%. Regardless of the value of I^2^ test, we a priori planned to perform sub-group analysis of the primary outcome according to severity of enrolled patients, accordingly, included studies were divided into two subgroups, based on whether endotracheal intubation and ICU admission was an exclusion criteria or it was allowed during enrollment, another subgroup analysis was planned based on TCZ dose (single or multiple). Furthermore, we planned a meta-regression analysis for the primary outcome based on the following criteria: patients’ severity (dichotomous), TCZ dose (dichotomous single or multiple), and number of recruited patients (continuous). We presented log odds ratio (LogOR), 95% CI, and p values of the meta-regression.

As a sensitivity test for the primary outcome we also presented RR of the less conservative (narrower CI) fixed effect model ^[15]^. All statistical tests and graphs were generated using STATA 14 software (StataCorp. 2015. *Stata Statistical Software: Release 14*. College Station, TX: StataCorp LP) and Review Manager (RevMan) [Computer program]. Version 5.3. Copenhagen: The Nordic Cochrane Centre, the Cochrane Collaboration, 2014.

The protocol of this study was reviewed and approved by the local institutional review board at King Saud Medical City, Riyadh, Saudi Arabia, under the registration number: H1R1 – 22 – Feb 21-01.

## Results

Our systematic search in PUBMED, EMBASE, and medXriv databases resulted in the inclusion of 9 articles ^[11, 12,16-22]^. Eight articles were duplicates between PUBMED and EMBASE, while one article ^[12]^ was unique to MedXriv, figure 1 shows the flow diagram of studies’ inclusion (details of excluded studies provided in table S1, supplementary file). All studies were randomized clinical trials according to our inclusion criteria with only three double blind studies ^[11, 17, 21]^. Three studies ^[17,21, 22]^ were multinational studies, the remaining studies although performed in a single country were all multicenter. The included studies recruited a total of 6326 patients, of whom 3272 patients were randomized to the intervention, and 3054 patients were randomized to control group. Included studies had multiple discrepancies among them, the most striking was the mechanical ventilation status of recruited patients, as four studies excluded patients if they were mechanically ventilated ^[11,16-18]^, whereas in the other five studies patients could be enrolled if they were mechanically ventilated ^[12,19-22]^.Only two studies provided TCZ as a single dose ^[11, 19]^, in the rest of the studies a second dose could be given if the patients were not improving clinically. It’s worth noting that the primary outcome of our review (28 – 30 day all-cause mortality) was the primary outcome for only one study ^[12]^ (table 1: Details of included studies). Publication bias assessment was done for the nine included studies which all contributed to the primary outcome, and despite fairly visually apparent lack of studies on the left (TCZ) side and apparent asymmetry, the p value of Egger’s test was insignificant (p = 0.201), indicating that there is no effect of small studies, however, trim and fill test indicated that 2 studies were missing on the left side (Figure S1, supplementary file).

**Figure 1:**
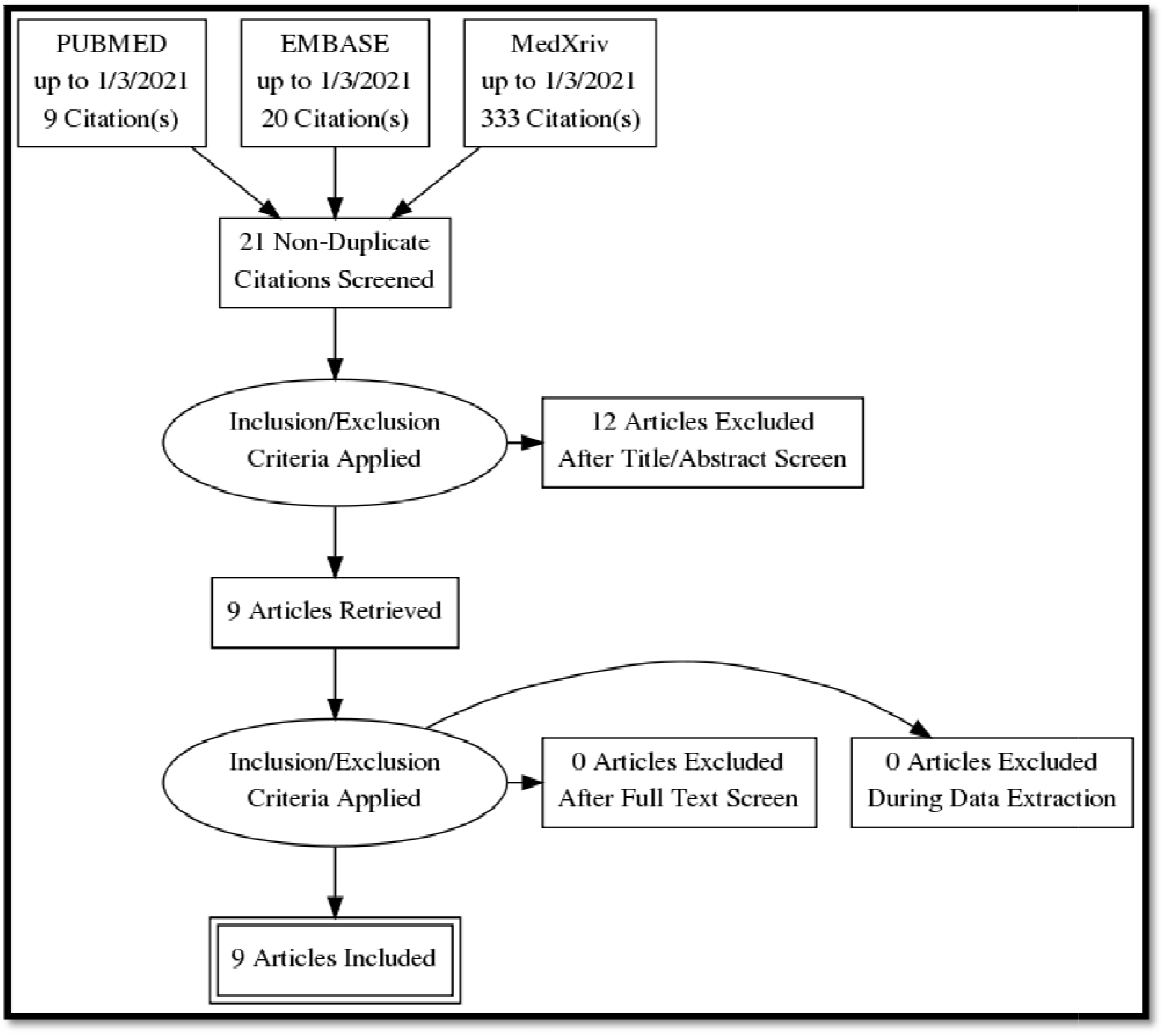
Studies inclusion flow diagram:

**Table 1:** Characteristics of included studies:

| Study | Inclusion Criteria | Exclusion Criteria | Tocilizumab Dose | Control | Primary outcome | Blinding | Place of trial |
| --- | --- | --- | --- | --- | --- | --- | --- |
| Stone [11] | 19 – 85 years, need O<sub>2</sub> to keep SpO<sub>2</sub> > 92% | O<sub>2</sub> > 10 L/min | 8 mg / kg (max 800 mg, single dose) | Standard of care | Intubation or death as time – event analysis. | Double Blind | USA, multicenter. |
| Horby [12] | SpO<sub>2</sub> < 92% on room air. Could be ventilated | hypersensitivity to tocilizumab, active tuberculosis infection or clear evidence of active bacterial, fungal, viral, or other infection. | 800 mg if weight >90kg; 600 mg if weight >65 and ≤90 kg; 400 mg if weight >40 and ≤65 kg; and 8mg/kg if weight ≤40 kg).<br>A second dose could be given 12 to 24 hours later. | Standard of care | All cause 28-day mortality | Open Label | U.K. multicenter. |
| Hermine [16] | 18 plus, moderate pneumonia (3L/min and 5L/min of oxygen to maintain SpO<sub>2</sub> >97%)<br>Severe: Respiratory distress (≥ 30 breaths/min); Oxygen saturation ≤93% at rest in ambient air; or Oxygen saturation ≤97% with O<sub>2</sub> > 5L/min.<br>PaO<sub>2</sub>/FiO<sub>2</sub> ≤ 300mmHg | high-flow oxygen(HFO) more than 15 L/minO<sub>2</sub>),<br>noninvasive ventilation(NIV) or mechanical ventilation (MV) and patients with critical pneumonia defined as WHO-CPS score of 6 or more (ie, with HFO,NIV, or MV).<br>ICU admission | intravenously (IV) at 8 mg/kg on day 1.<br>additional fixed dose 400 mg IV, on day 3 was recommended if oxygen requirement was not decreased by more than 50%, but decision was left to the treating physician. | Standard of care | the proportion of patients dead or needing noninvasive or mechanical ventilation on day 4 | Open label | France, multicenter. |
| Salama [17] | 18 and older<br>SpO<sub>2</sub> < 94% while breathing ambient air | continuous positive airway pressure, bilevel positive airway pressure, or mechanical ventilation. | intravenous 8 mg per kilogram of body weight, to a maximum of 800 mg per dose)<br>clinical signs or symptoms worsened or did not improve, an additional infusion could be administered 8 to 24 hours after the first one. | Standard of care | mechanical ventilation / ECMO /or death by day 28. | Double Blind | USA, Peru, Brazil, Kenya, South Africa, Mexico. |
| Salvarani [18] | 18 or more, P/F ratio 200 – 300, allowed to receive oxygen therapy with Venturi mask or high-flow nasal cannula | P/F ratio < 200, MV or NIMV, ICU admission, Shock, Heart / kidney failure | intravenously 8mg/kg up to a maximum of 800 mg within 8 hours of randomization, followed by a second dose after 12 hours. | Standard of care. | Clinical worsening within 14 (Admission to ICU with mechanical ventilation<br>Death from any cause | Open Label | Italy, multicenter |
|  |  |  |  |  | PaO<sub>2</sub>/FIO<sub>2</sub> ratio less than 150 |  |  |
| Veiga [19] | 18 or more, suppl O<sub>2</sub> to keep SpO<sub>2</sub> > 93% OR MV for less than 24 hours, | active uncontrolled infection, raised aspartate aminotransferase or alanine aminotransferase levels greater than five times the upper limit of normal, and renal disease with an estimated glomerular filtration of <30 mL/min/1.72 m<sup>2</sup>. | single intravenous infusion of 8 mg/kg) | Standard of care. | clinical status at 15 days evaluated with the use of a seven level ordinal scale, | Open label | Brazil, multicenter. |
| Zhao [20] | more than 18 years old, could be mechanically ventilated | Allergic to tocilizumab; Pregnant or lactating ALT or AST > 5 times of upper limit of normal; active hepatitis, tuberculosis, and definite bacterial or fungal infections; | 4– 8 mg/kg (recommended 400 mg)<br>For patients with fever, if there was still fever within 24 h after the first used, it should be used once more (the dose was the same as before). | Favipiravir | cumulative lung lesion remission rate (lung CT examination indicated absorption of lung inflammation). | Open label | China, multicenter |
| Rosas [21] | ≥18 years of age oxygen saturation of 93% or less or a P/F ratio of less than 300 mm Hg. Could be mechanically ventilated. | Eminent death with 24 hours, tuberculosis or a bacterial, fungal, or viral infection other than SARS-CoV-2. | 8 mg per kilogram of body weight, with a maximum dose of 800 mg<br>If clinical signs or symptoms did not improve or worsened, a second infusion of tocilizumab or placebo could be administered 8 to 24 hours after the first dose. | Standard of care | clinical status at day 28, as assessed on the seven-category ordinal scale. | Double Blind | Canada, Denmark, France, Germany, Italy, the Netherlands, Spain, the United Kingdom, and the United States |
| Gordon [22] | >or equal 18 admitted to ICU receiving respiratory or cardiovascular organ support | Imminent death | 8mg/kg of actual body weight (up to a maximum of 800mg)<br>could be repeated 12-24 hours later at the discretion of the treating clinician. | Standard of care | Respiratory and cardiovascular organ support free days at day 21. | Open label | International platform |

Our assessment of RoB of the included studies was low in five domains for all studies, whereas the domain of random sequence generation (selection bias) was assessed as unclear in three studies ^[12,18,19]^ since these studies didn’t report the number of screened patients for eligibility, RoB was also deemed unclear in the “Other” domain for one study, in view of the significant involvement of the sponsor in the study’s design, conduct, data collection, and analysis ^[17]^. This means that the overall RoB was 100% low in 5 domains, 67% low in the selection bias, and 89% low in “Other” domain (figure 2 a and b).

**Figure 2:**
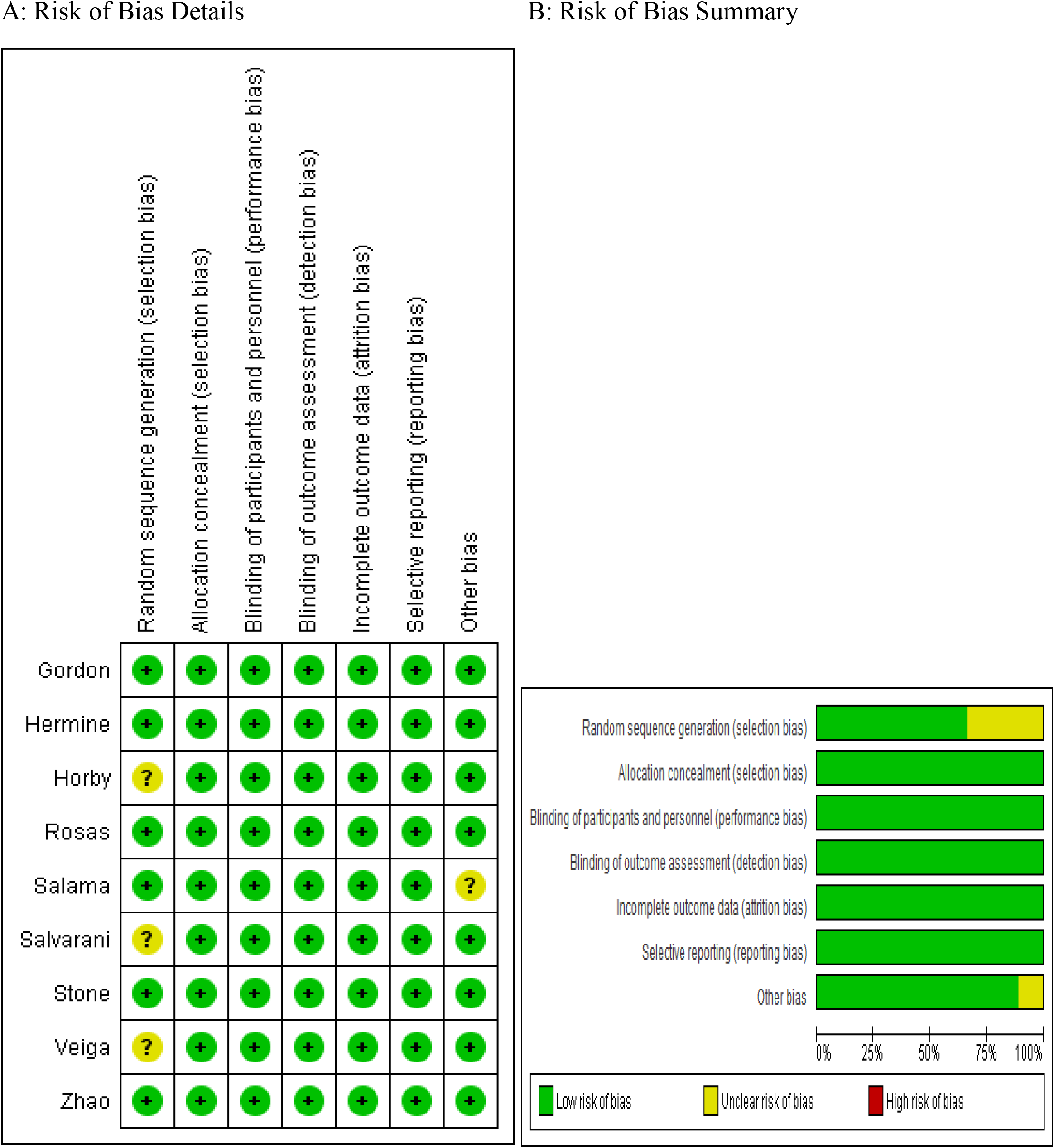
Risk of Bias details and summary:

### Primary outcome: 28-30 day all-cause mortality

In the intervention (TCZ) pooled arm 810 incidences of death occurred within 28-30 days follow up period out of a total of 3272 patients, whereas in the pooled control arm 895 incidences of death occurred within the same follow up period out of 3054 patients. Intuitively, this result indicates lower mortality in the TCZ arm, however, the result just missed statistical significance in the random effects model with RR = 0.9 (95% CI: 0.8 – 1.01; p = 0.09). (Figure 3). Heterogeneity among the studies included in the primary outcome was very low at I^2^ value of only 9%, this low heterogeneity is also reflected in an insignificant p value (0.36) of chi square test of heterogeneity. However, this statistically insignificant impact of TCZ on 28-30 day mortality was not robust in our predefined sensitivity test of fixed effect. In the fixed effect model RR = 0.9 (95% CI: 0.83 – 0.97; p = 0.008).

**Figure 3:**
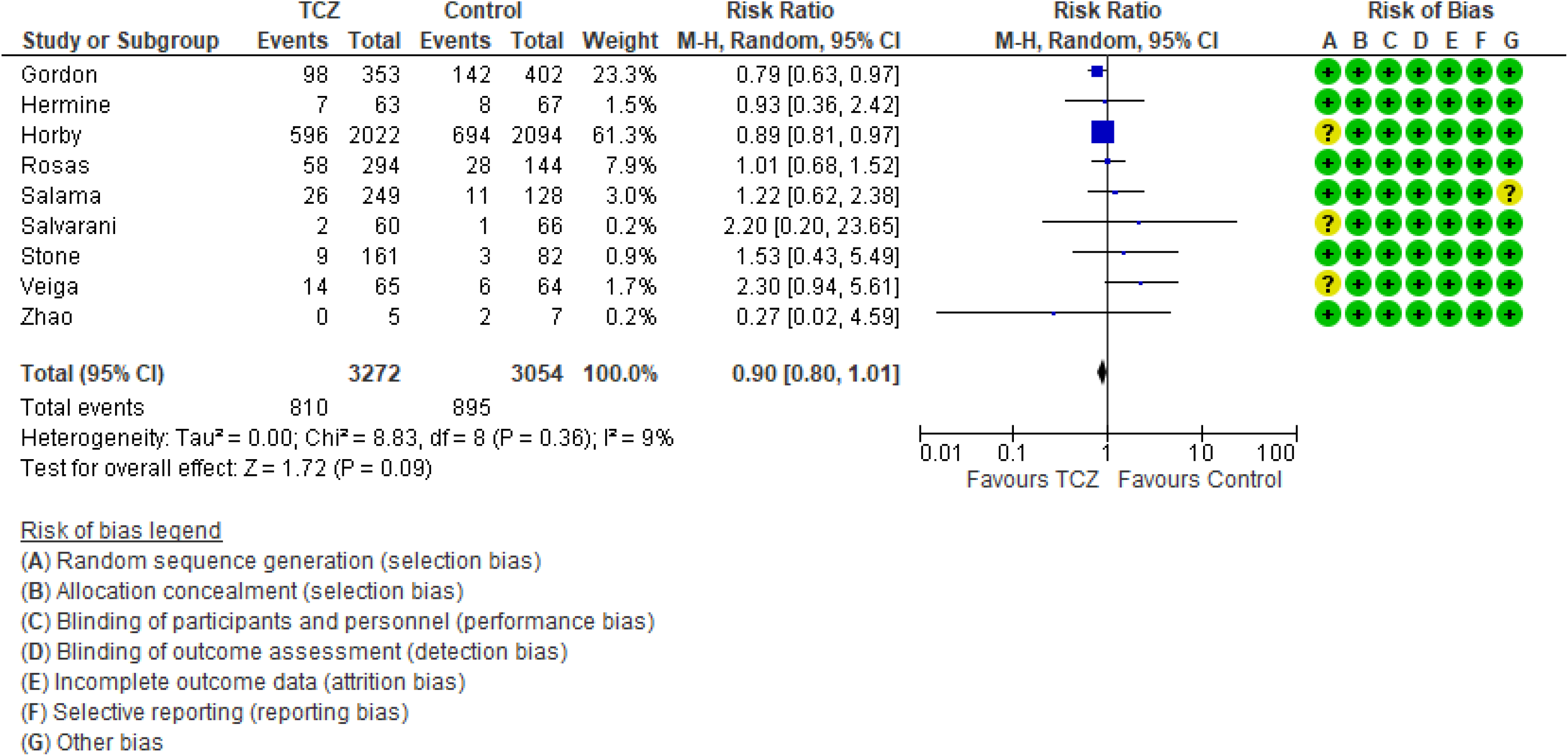
28 – 30 day all-cause mortality forest plot: Random effects model.

Interestingly, the primary outcome had very low heterogeneity despite the fact that different populations were included, different doses of TCZ given, and substantially variable sample size for each study. Accordingly, we decided to proceed as planned with our sub-group and meta-regression analyses. Subgroup analyses based on the number of TCZ doses indicates that in the two studies allowing only one dose RR = 2.011 (95% CI: 0.97 – 4.2; p = 0.06) that is to say there was no statistical difference although clinically the effect was in favor of the control group. On the contrary, in the subgroup of studies that allowed more than one dose of TCZ, there was a statistically significant reduction of mortality in the TCZ group (RR = 0.9, 95% CI: 0.81 – 0.96; p = 0.003). The second predefined subgroup analysis was based on criteria of inclusion, in the subgroup of studies not recruiting mechanically ventilated patients there was no difference between both groups with regards to mortality (RR= 1.2, 95% CI: 0.7 – 2; p = 0.4). Similarly, in the subgroup allowing recruitment of mechanically ventilated patients there was no difference in mortality as well (RR = 0.9, 95% CI: 0.8 – 1.1, p = 0.24). The supplementary file has more details in figures S3 – S6).

### Meta regression

Three predefined variables were used to perform the meta-regression, number of TCZ doses (as a binary variable), whether recruitment of mechanically ventilated patients was allowed (as a binary variable), and the total number of recruited patients in each study (as a continuous variable). The only variable with statistical significance was the use of more than one dose of TCZ, as it showed a significant reduction of mortality. For this variable, the coefficient was −0.91 (95% CI: −0.04 to −1.77; p = 0.04). The other two variables were not statistically significant (Details in table S2 and figures S7 – S9 in supplementary file).

### Secondary outcomes

The first secondary outcome was the combined outcome of either death, intubation, or admission to ICU (we collectively call this outcome: Clinical worsening). Only five out of the included nine studies contributed toward this outcome (figure 4) including 2352 patients in the TCZ arm and 2107 patients in the control arm. There was a statistically significant reduction of this composite outcome with the administration of TCZ, as RR = 0.83 (95% CI: 0.76 – 90; p < 0.001), there was no heterogeneity detected between studies contributing to this outcome (I^2^ = 0%, and chi square test of heterogeneity had an insignificant p value of 0.45).

**Figure 4:**
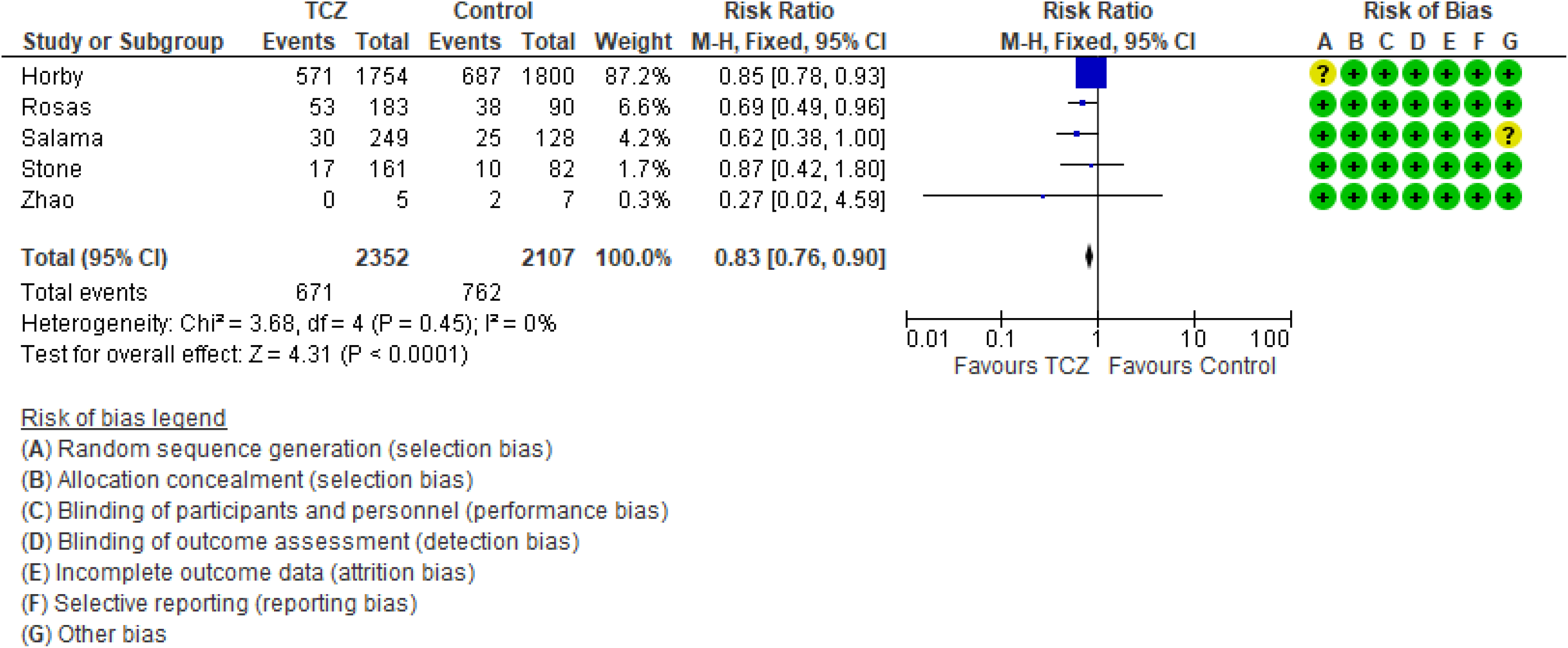
28 – 30 day Mortality / Intubation / ICU admission forest plot: Random effects model.

The second secondary outcome was the safety outcome of super-added infection, six studies contributed to this outcome, that have reported the incidence of infection out of a total of 892 patients who received TCZ, and 551 patients in the control arm. This outcome showed statistically insignificant RR between both arms (RR = 0.77, 95% CI: 0.51 – 1.19; p = 0.24). Figure 5 shows the forest plot of the superadded infection outcome.

**Figure 5:**
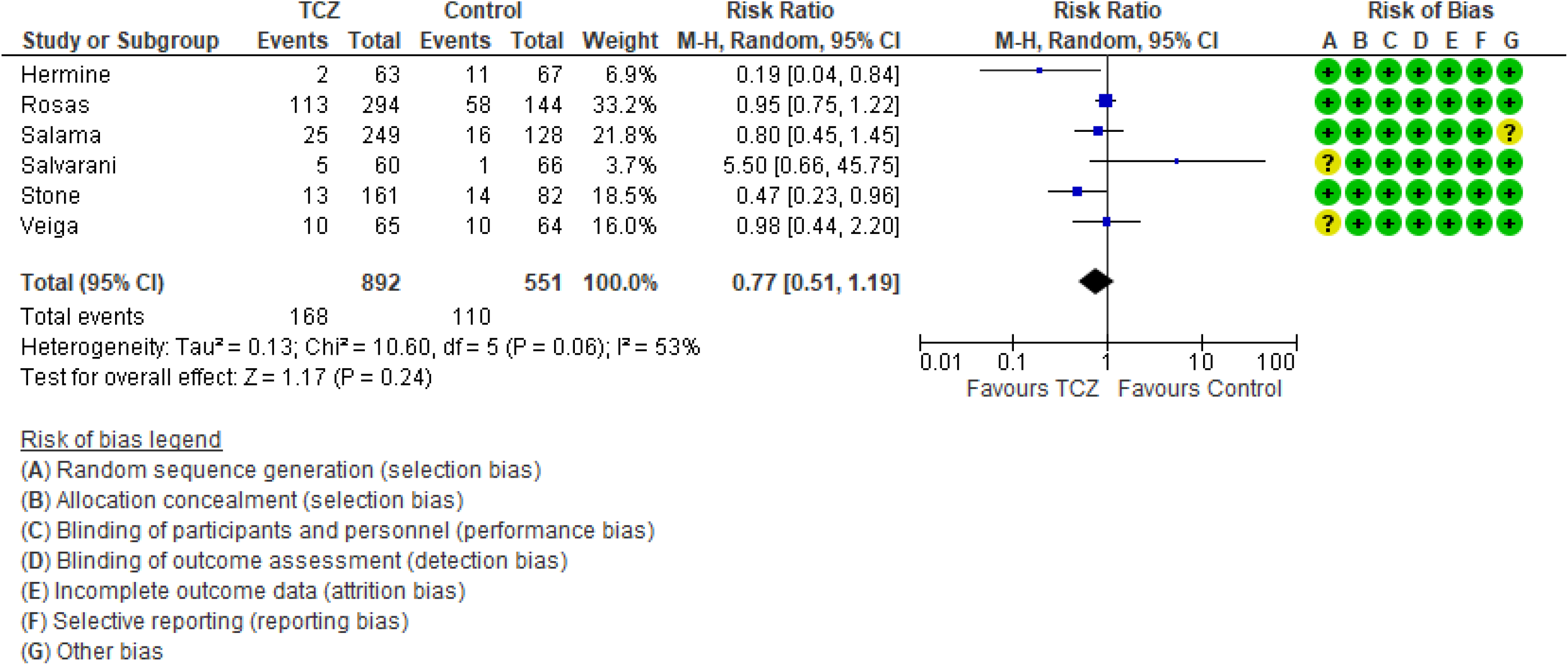
Superadded infection forest plot: Random effects model.

### Certainty of evidence

We utilized the GRADE approach to evaluate the certainty of evidence for our review’s three outcomes. The primary outcome of 28-30 day all-cause mortality and the secondary outcome of composite death / intubation / ICU admission were both downgraded once to moderate in view of their relatively wide 95% CI and the inclusion of several small studies. The secondary outcome of super-added infection was downgraded twice due to the same reason as the previous two in addition to high heterogeneity with I^2^ = 53%. Table S3 in supplementary file provides further details.

## Discussion

This systematic literature review and meta-analysis may not be unique in investigating the effects of TCZ on the outcomes of COVID-19 hospitalized patients, it is –however-the most updated in terms of inclusion of RCTs on the topic, other reviews ^[23]^ had fewer published RCTs available to them at the time of their publication, hence, this review could be considered as an update of previously published information. In this review we included a total of 6326 patients from 9 RCTs that compared receiving TCZ plus standard of care (3272 patients) to standard of care alone (3054 patients) in the management of hospitalized COVID-19 pneumonia. The intuition of a presumed benefit of Il-6 receptor antagonists arisen early in the COVID-19 era in view of similarities of the pathophysiology of COVID-19 pneumonia to other conditions associated with cytokine release syndrome, such as haemophagocytic lymphohistiocytosis and macrophage activation syndrome ^[24]^ and that this cytokine release syndrome is responsible for the multi-organ failure commonly associated with COVID-19 infection, particularly its severe forms ^[24]^, accordingly, the use of humanized monoclonal antibodies against IL-6 receptors such as TCZ may mitigate the dysregulated host immune response in COVID-19 infection, and subsequently avoid associated lung tissue damage ^[5]^.

These benefits of TCZ particularly on mortality were demonstrated by several observational studies as well as reviews including observational studies ^[7,10]^, however, such results are questionable in view of the inherent limitations of observational studies in terms of design, in addition to patients’ severity and clinical condition variations ^[23]^, furthermore, other reviews failed to demonstrate such benefits, the review by Lan SH et al ^[25]^ concluded no additional benefits of TCZ on mortality, mechanical ventilation, and ICU admission, although these results should also be looked at cautiously, in view of the substantial heterogeneity in all three outcomes, and of course the observational nature of the included studies. What was agreed upon by almost all the early studies and reviews was the need for well-designed randomized clinical trials.

In our review the outcome of short term (28 – 30 day) mortality showed no statistical significance of TCZ, however, this result should be meticulously examined, as it only reports statistical significance, while overlooking potential clinical benefit, the RR was found to be 0.9, however, the 95% CI was 0.8 – 1.01, with an overall effect p value of 0.09. While statistically insignificant, we should understand that the significance was only missed by 0.01 in the 95% CI, more importantly, the result should not be interpreted as lack of effect, but rather as not enough evidence to reject the null hypothesis, and consequently, a clinically meaningful effect can’t just be ruled out based solely on statistical results ^[26]^. Furthermore, the statistically insignificant result of our primary outcome didn’t withstand the sensitivity test of fixed effect model (in fixed effect model RR = 0.9, and 95% CI ranged between 0.83 and 0.97; p = 0.008), although the random effects model is the most valid model of the two in view of differences between studies at least in terms of patients’ conditions and doses of TCZ ^[27]^. What further strengthens the impression that there could be a meaningful clinical effect of TCZ on mortality despite a statistically insignificant overall effect is the fact that the trim and fill funnel plot (figure S1, supplementary file) indicates two missing studies in favor of TCZ (the left side), that is to say our result may be underpowered. This is supported by the findings of a similar review ^[23]^ in which only 5 RCTs were included, and the overall effect on mortality was highly insignificant (RR = 1.09, 95% CI: 0.8 – 1.49; p = 0.57), in the study by RECOVERY Collaborative group ^[12]^ a meta-analysis section of previously published RCTs was included, in that section the addition of three more RCTs yielded a statistically significant result in favor of TCZ (RR = 0.87, 95% CI: 0.79 – 0.96; p = 0.005). While the addition of a ninth small RCT in our review ^[20]^ that showed no difference in mortality resulted in widening of the 95% CI to miss statistical significance.

Despite the fact that heterogeneity was substantially low (9%), subgroup analysis a priori determined indicated both a statistically significant and a clinically meaningful reduction of mortality among patients treated with more than one dose of TCZ, regardless of whether critically ill (mechanically ventilated) patients were recruited or not (figure S4, supplementary file). This finding was confirmed in our meta regression analysis, where doses of TCZ (strictly one dose versus possible second dose) was the only significant predictor of lower mortality, in contrast to the severity of recruited patients which proved insignificant in both subgroup and meta regression analyses.

Both of our secondary objectives showed results in favor of treatment with TCZ both statistically and clinically, RR of composite outcome of death, mechanical ventilation, or ICU admission was lower in the group treated with TCZ, while there was no difference in the incidence of super-added infection as a safety measure. Notably, those two secondary outcomes included only five and six studies respectively, indicating under-power.

It is worth mentioning that the certainty of evidence of the primary outcome, and the secondary composite outcome of death, intubation, or ICU admission were both downgraded to “Moderate” in view of imprecision due to the inclusion of small sized studies with few events, and wide confidence intervals. The secondary outcome of super-added infection was downgraded twice to “Low” because of imprecision (previously described) and inconsistency with an I^2^ test of heterogeneity of 53%.

## Conclusion

We conclude that moderate certainty of evidence suggest no statistically significant improvement of 28-30 day all-cause mortality of hospitalized COVID-19 patients treated with TCZ, although there may be clinically important value. Moderate certainty of evidence suggest lowered relative risk of a composite outcome of death or clinical deterioration, while, low grade evidence indicate no increase in the risk of super-added infection associated with TCZ treatment. A protocol allowing two doses of TCZ shows evidence of improved mortality as compared to a strictly single dose protocol.

## Data Availability

Data available upon request from the corresponding author by e-mail.

## Strengths and Limitations

Our review included only RCTs and all available RCTs on the topic to our best knowledge, they were all well-designed with very low levels of RoB. We utilized rigorous statistical methods of meta-analysis, subgroup analysis, and meta-regression, despite low between studies statistical heterogeneity, since clinical heterogeneity was clearly evident at least with regards to study design, blinding, and inclusion criteria.

However, our review is subject to several limitations as well, we included data from one preliminary report of a study available at medRxiv but is still not officially published and accordingly not peer reviewed, although with a high level of validity since it was a very well designed study by a highly trusted group. We also included a small study from China that recruited a limited number of patients (12 patients), and this particular study showed no difference in mortality, but had a very wide 95% CI, that had an obvious impact on the overall effect in the primary outcome. Our review examined three outcomes only, the outcomes that appeared to be most patient centered, however, many more outcomes were studied by the included articles, yet we didn’t include them as they were not consistent in the included articles, and investigating them would have resulted in a small number of studies in each outcome. Finally, the primary outcome of our review was in fact the primary outcome of only one included study, in other words, eight out of nine studies were not sufficiently powered to detect the impact of TCZ on short term mortality.

## Acknowledgment

The authors would like to extend their gratitude to:

Mrs. Huda Mhawish, nursing supervisor of ICU at King Saud Medical City.

Ms. Katrina Baguisa, study coordinator.

Ms. Alva M. Alcazar. Study coordinator.

