## Supplementary file for "Tocilizumab Effect in COVID-19 Hospitalized Patients: A Systematic Review and Meta-Analysis of Randomized Control Trials"

1- Detailed PUBMED search strategy.
2- Table S1: Excluded studies.
3- Figure S1: Publication bias funnel plot.
4- Figure S2: Fixed effect model of primary outcome.
5- Figure S3: Subgroup analyses of primary outcome: Single dose TCZ.
6- Figure S4: Subgroup analyses of primary outcome: More than one dose TCZ.
7- Figure S5: Subgroup analyses of primary outcome: Mechanical ventilation not allowed.
8- Figure S6: Subgroup analyses of primary outcome: Mechanical Ventilation allowed.
9- Table S2: Results of Meta regression.
10- Figure S7: Bubble Plot: Number of patients.
11- Figure S8: Bubble Plot: Mechanical Ventilation.
12- Figure S9: Bubble plot: Doses of TCZ.
13- Table S3: Summary of findings and GRADE of evidence.

**Detailed PUBMED search strategy:**

(("covid 19"[All Fields] OR "covid 19"[MeSH Terms] OR "covid 19 vaccines"[All Fields] OR "covid 19 vaccines"[MeSH Terms] OR "covid 19 serotherapy"[All Fields] OR "covid 19 serotherapy"[Supplementary Concept] OR "covid 19 nucleic acid testing"[All Fields] OR "covid 19 nucleic acid testing"[MeSH Terms] OR "covid 19 serological testing"[All Fields] OR "covid 19 serological testing"[MeSH Terms] OR "covid 19 testing"[All Fields] OR "covid 19 testing"[MeSH Terms] OR "sars cov 2"[All Fields] OR "sars cov 2"[MeSH Terms] OR "severe acute respiratory syndrome coronavirus 2"[All Fields] OR "ncov"[All Fields] OR "2019 ncov"[All Fields] OR (("coronavirus"[MeSH Terms] OR "coronavirus"[All Fields] OR "cov"[All Fields]) AND 2019/11/01:3000/12/31[Date - Publication]) OR ("sars cov 2"[MeSH Terms] OR "sars cov 2"[All Fields] OR "sars cov 2"[All Fields])) AND "randomized controlled trial"[Publication Type] AND (("tocilizumab"[Supplementary Concept] OR "tocilizumab"[All Fields]) AND "randomized controlled trial"[Publication Type])) AND (randomizedcontrolledtrial[Filter])

Table S1: Excluded studies after review of Abstract / Full test:

| Study Reference | Reason of exclusion |
| --- | --- |
| Andrew Tsai, et al. [Impact of tocilizumab administration on mortality in severe COVID-19](https://www.medrxiv.org/content/10.1101/2020.07.30.20114959v1). medRxiv 2020.07.30.20114959; doi: <https://doi.org/10.1101/2020.07.30.20114959> | COHORT |
| Garth W. Strohbehn, et al. [COVIDOSE: Low-dose tocilizumab in the treatment of Covid-19](https://www.medrxiv.org/content/10.1101/2020.07.20.20157503v1). medRxiv 2020.07.20.20157503; doi: <https://doi.org/10.1101/2020.07.20.20157503> | Historical Controls |
| Emily C Somers, et al. [Tocilizumab for treatment of mechanically ventilated patients with COVID-19](https://www.medrxiv.org/content/10.1101/2020.05.29.20117358v1). medRxiv 2020.05.29.20117358; doi: https://doi.org/10.1101/2020.05.29.20117358 | COHORT |
| Victor Carvalho, et al. [Effects of Tocilizumab in Critically Ill Patients With COVID-19: A Quasi-Experimental Study](https://www.medrxiv.org/content/10.1101/2020.07.13.20149328v1). medRxiv 2020.07.13.20149328; doi: https://doi.org/10.1101/2020.07.13.20149328 | Historical controls |
| Francesco Perrone, et al. [Tocilizumab for patients with COVID-19 pneumonia. The TOCIVID-19 prospective phase 2 trial](https://www.medrxiv.org/content/10.1101/2020.06.01.20119149v2). medRxiv 2020.06.01.20119149; doi: https://doi.org/10.1101/2020.06.01.20119149 | Single arm, not compared to standard. |
| Malgorzata Mikulska, et al. [Tocilizumab and steroid treatment in patients with COVID-19 pneumonia](https://www.medrxiv.org/content/10.1101/2020.06.22.20133413v1). medRxiv 2020.06.22.20133413; doi: https://doi.org/10.1101/2020.06.22.20133413 | Observational |
| Manuel Rubio-Rivas, et al. [Beneficial Effect of Corticosteroids in Preventing Mortality in Patients Receiving Tocilizumab to Treat Severe COVID-19 Illness](https://www.medrxiv.org/content/10.1101/2020.08.31.20182428v1)  medRxiv 2020.08.31.20182428; doi: https://doi.org/10.1101/2020.08.31.20182428 | Observational |
| Nafisa Wadud. Et al. [Improved survival outcome in SARs-CoV-2 (COVID-19) Acute Respiratory Distress Syndrome patients with Tocilizumab administration](https://www.medrxiv.org/content/10.1101/2020.05.13.20100081v1). medRxiv 2020.05.13.20100081; doi: https://doi.org/10.1101/2020.05.13.20100081 | Retrospective |
| Martínez-Sanz J, et al. Effects of tocilizumab on mortality in hospitalized patients with COVID-19: a multicentre cohort study. Clin Microbiol Infect. 2021;27(2):238-243. doi:10.1016/j.cmi.2020.09.021 | COHORT |
| Tian J, et al. Repurposed Tocilizumab in Patients with Severe COVID-19. J Immunol. 2021;206(3):599-606. doi:10.4049/jimmunol.2000981 | Retrospective observational. |
| Pomponio G, et al. Tocilizumab in COVID-19 interstitial pneumonia. J Intern Med. 2021 Jan 29. doi: 10.1111/joim.13231. | Single arm, not compared to standard. |
| Jordan SC, et al. Compassionate Use of Tocilizumab for Treatment of SARS-CoV-2 Pneumonia. Clin Infect Dis. 2020 Dec 15;71(12):3168-3173. doi: 10.1093/cid/ciaa812. | Single arm, not compared to standard. |

Figure S1: Publication Bias funnel plot.

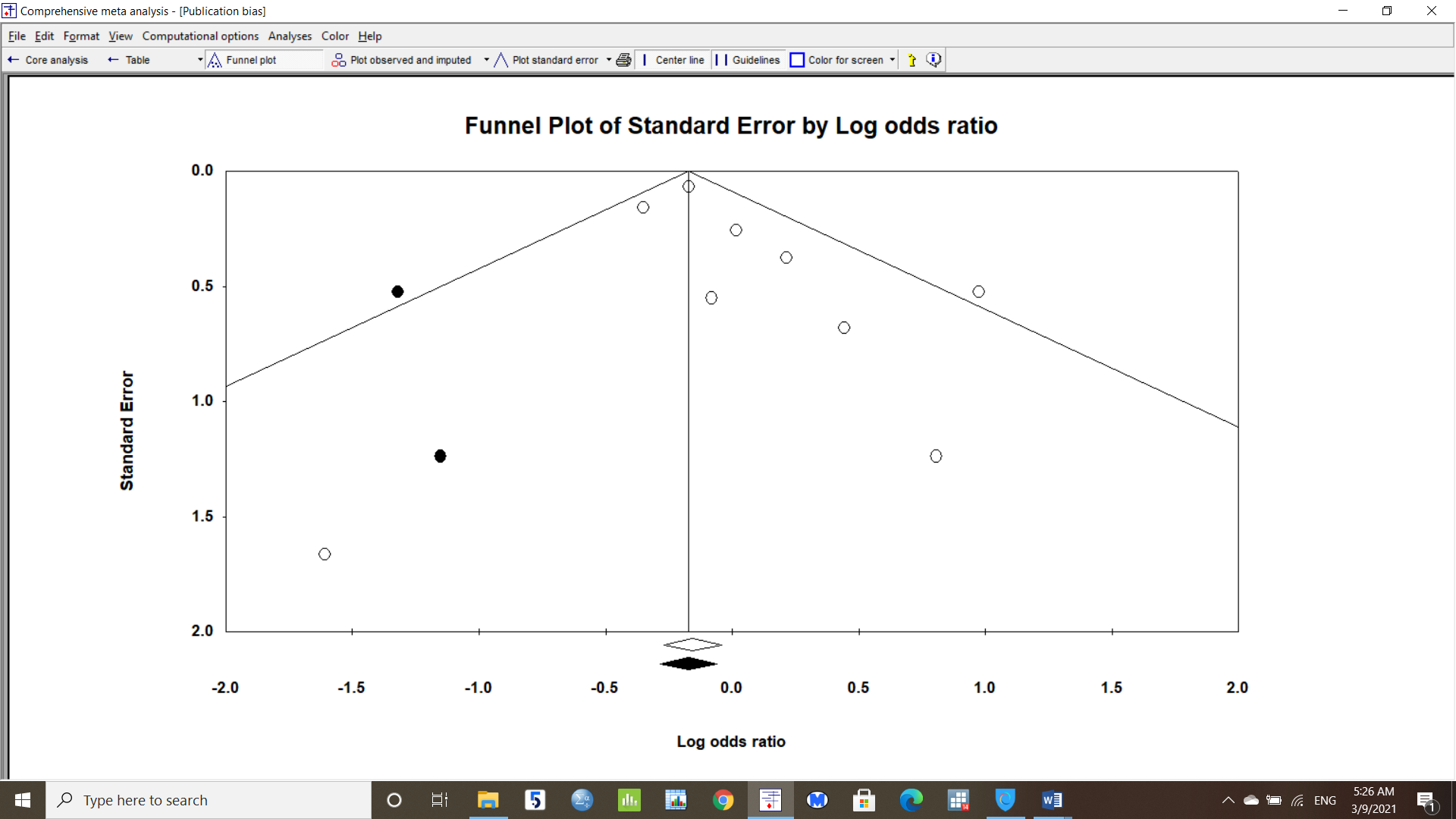

Included studies

Missing studies

Figure S2: Fixed effect model of Primary outcome:

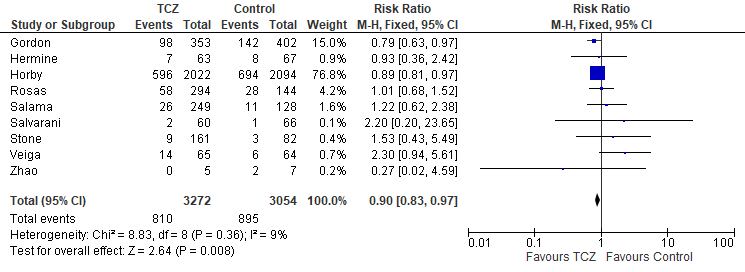

Figure S3: Primary outcome subgroup analysis: Single dose TCZ:

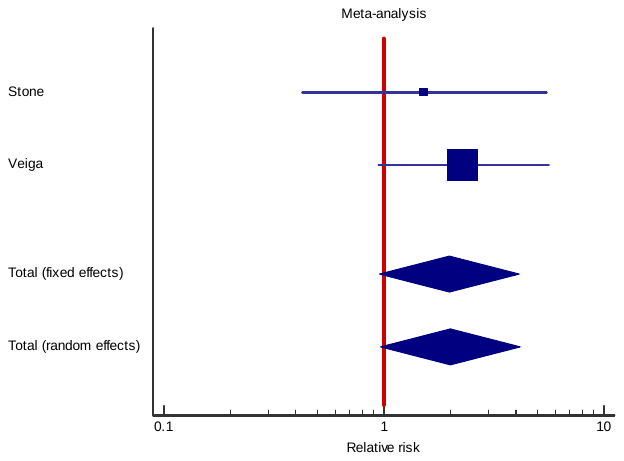

Random Effects: RR = 2.011 (95% CI: 0.97 – 4.2; p 0.06)

Figure S4: Primary outcome subgroup analysis: More than one dose of TCZ:

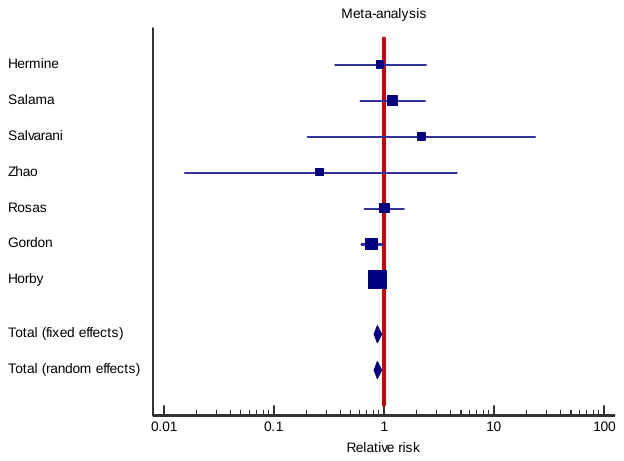

Random effects: RR = 0.9 (95% CI: 0.81 – 0.96; p = 0.003)

Figure S5: Primary outcome subgroup analysis: Mechanical ventilation not allowed upon recruitment:

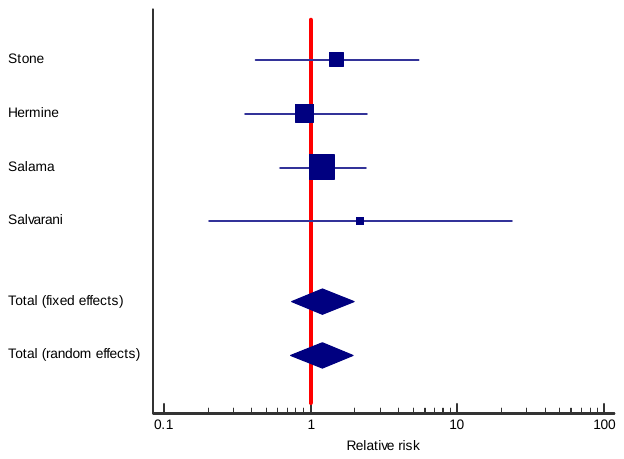

Random effects: RR = 1.2 (95% CI: 0.7 – 2, p =0.4)

Figure S6: Primary outcome subgroup analysis: Mechanical ventilation allowed during recruitment:

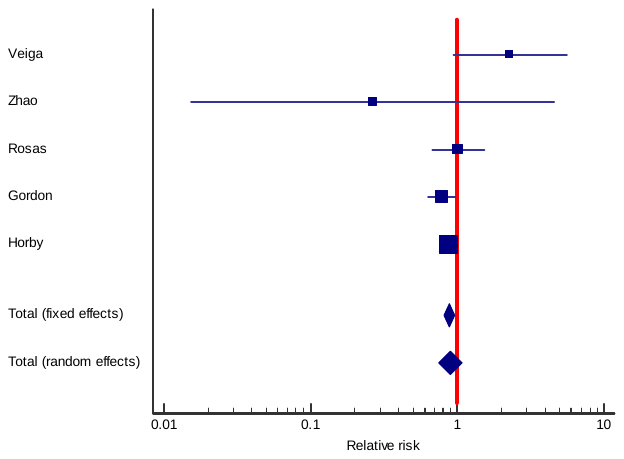

Random Effects: RR = 0.9 (95% CI: 0.8 – 1.1, p = 0.24)

Table S2: Results of Meta regression:

| **Covariate** | **Coefficient** | **Standard Error** | **95% Lower** | **95% Upper** | **Z value** | **P value (2 sided)** |
| --- | --- | --- | --- | --- | --- | --- |
| **Intercept** | **0.9609** | **0.4585** | **0.0623** | **1.8594** | **2.1** | **0.04** |
| **Total number of patients** | **0.00** | **0.00** | **-0.0001** | **0.0001** | **0.39** | **0.7** |
| **Mechanical ventilation (Allowed)** | **-0.2959** | **0.3131** | **-0.9096** | **0.3177** | **-0.95** | **0.3** |
| **TCZ Doses (two)** | **-0.9069** | **0.4409** | **-1.7711** | **-0.0427** | **-2.06** | **0.04** |

Figure S7: Bubble Plot: Number of patients:

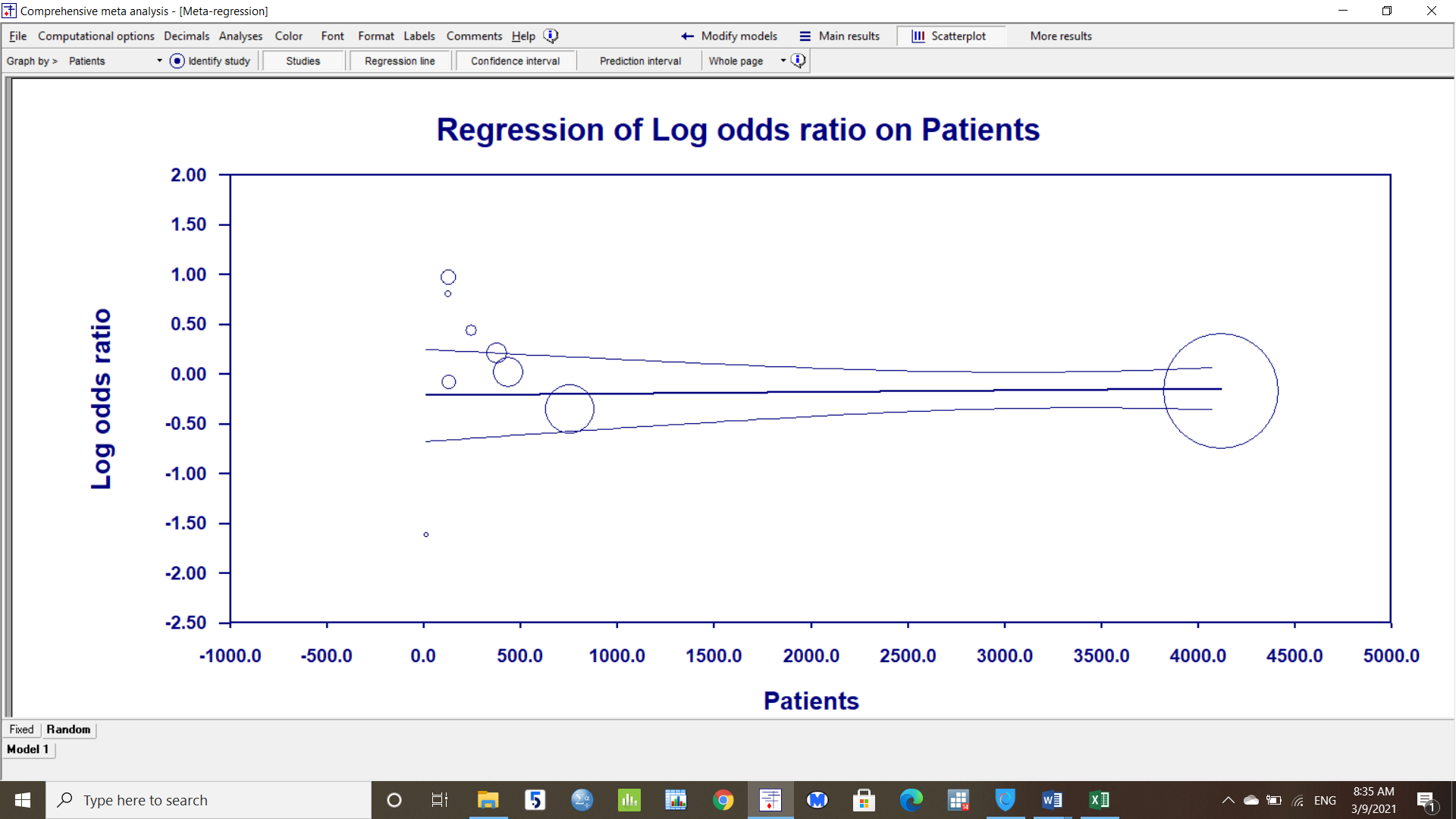

Figure S8: Bubble Plot: Mechanical Ventilation during recruitment.

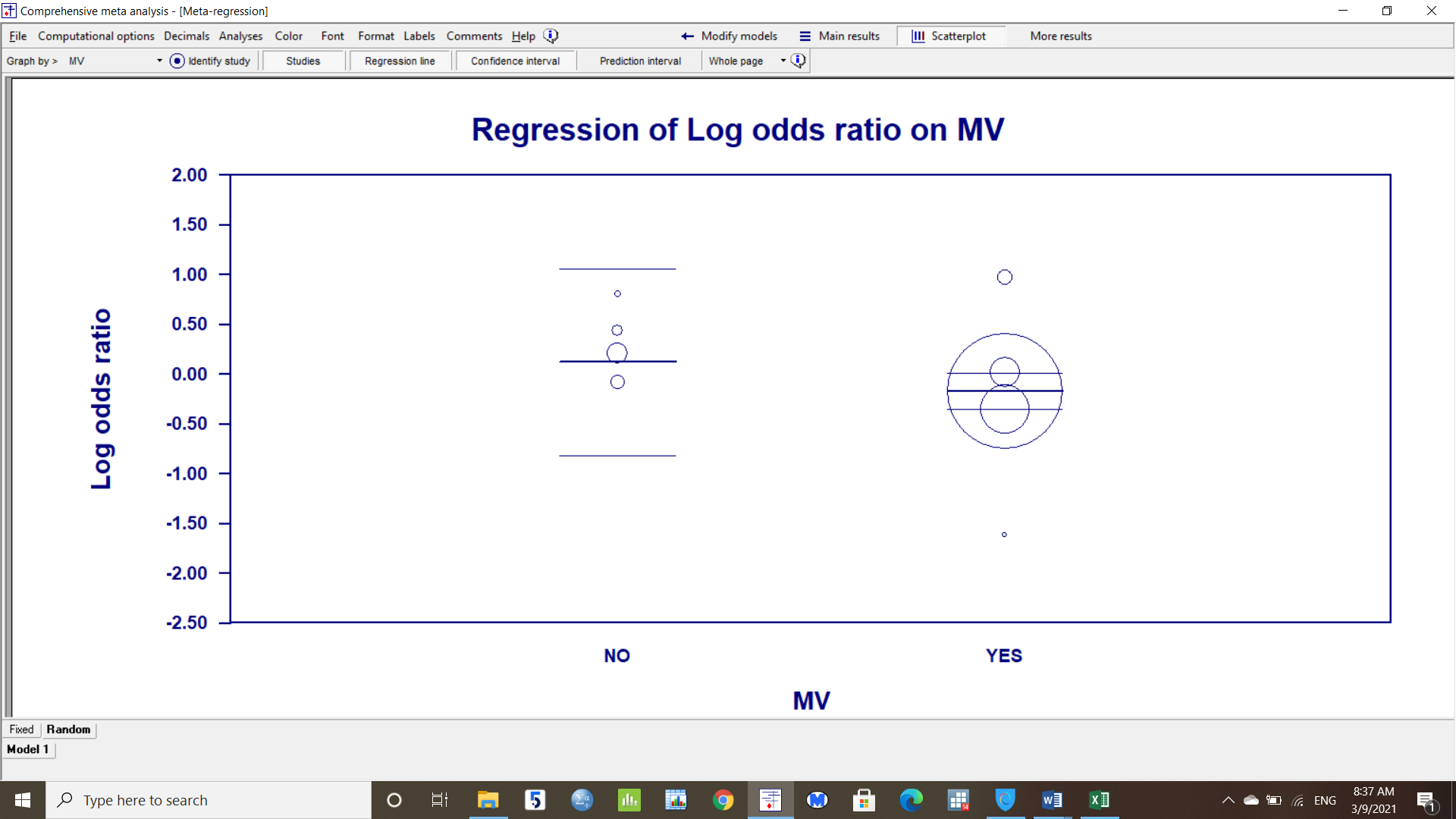

Figure S9: Bubble plot: Doses of TCZ:

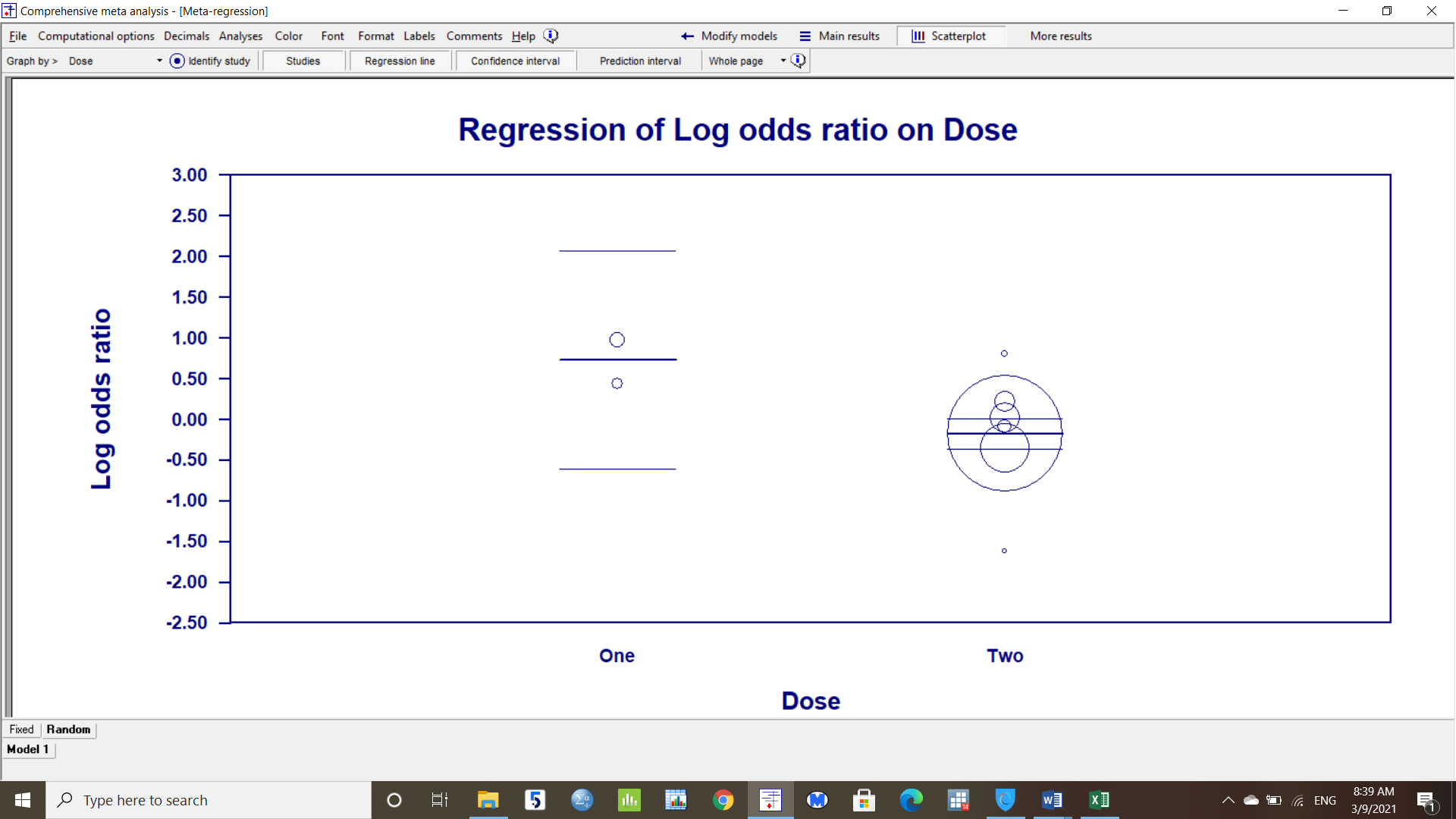

Table S3: Summary of findings table and GRADE evaluation:

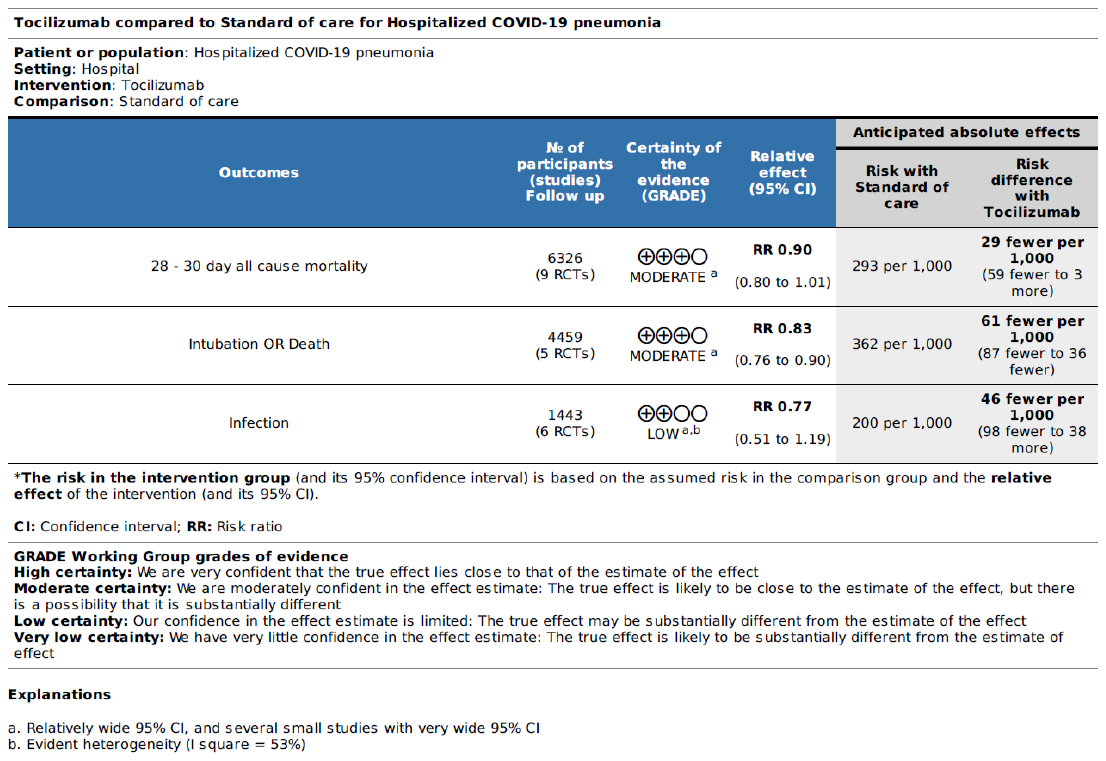
